# Successful strategies that address gender-related barriers and promote bodily autonomy within efforts to scale up and sustain post-pregnancy contraception: A scoping review

**DOI:** 10.1101/2024.06.21.24309318

**Authors:** Arachu Castro, Rita Kabra, Anna Coates, James Kiarie

## Abstract

**Introduction:** Acknowledging the integral role of bodily autonomy in advancing gender equality, our study aimed to assess the extent to which strategies used in postpartum and post-abortion contraception have effectively equipped women, girls, and gender-diverse individuals with the tools, knowledge, and resources required to make autonomous decisions that align with their diverse life experiences.

**Methods:** We conducted a scoping review using the databases PubMed, EBSCOhost, EMBASE, and SciSpace. We included implementation, evaluation, and experimental studies published in any language between 2013 and 2023 and excluded studies not meeting these criteria. We used a WHO scale to determine the level of gender responsiveness.

**Results:** We found 30 implementation, evaluation, and experimental studies published in any language between 2013 and 2023. We categorized the strategies as gender-transformative (4 studies), gender-specific (24 studies), and gender-sensitive (2 studies). None of the studies reported strategies hindering reproductive health and rights. All strategies involved women and girls, and none explicitly targeted gender-diverse people capable of childbearing.

**Conclusion:** Gender-transformative strategies shared a commitment to empowering women with the autonomy to make informed decisions about post-pregnancy contraception through a) delivering personalized counseling that respected each woman’s reproductive goals and ensured privacy during these discussions; b) integrating the cultural and familial context of women’s health decisions; and c) promoting a rights-based approach that prioritized informed consent and defended women’s reproductive rights. These interventions were delivered in contexts of quality improvement strategies integrating contraception services into the continuum of post-pregnancy care, offering women continuous access to information. Gender-specific strategies focused on informed contraceptive decision-making while recognizing the potential impact of gender dynamics on contraceptive use without actively challenging the underlying gender norms or power relations. Gender-sensitive strategies, although indicated gender awareness, did not address the process of informed contraceptive decision-making nor emphasize the provision of supportive environments that respect and enhance bodily autonomy.

**KEY MESSAGES:** *What is already known on this topic:* Previous scoping and systematic reviews have explored strategies to increase post-pregnancy contraception uptake globally. However, none have specifically focused on strategies that promote bodily autonomy while addressing gender-related barriers. Our study addresses this gap by providing a comprehensive understanding of such strategies and their impact on scaling up and sustaining post-pregnancy contraception.

*What this study adds:* This study provides new insights by being the first scoping review to focus on strategies promoting bodily autonomy in addressing gender-related barriers to scaling up and sustaining post-pregnancy contraception. The gender-transformative strategies reported in the studies shared a commitment to empowering women with the autonomy to make informed decisions about post-pregnancy contraception through a) delivering personalized counseling that respected each woman’s reproductive goals and ensured privacy during these discussions; b) integrating the cultural and familial context of women’s health decisions; and c) promoting a rights-based approach that prioritized informed consent and defended women’s reproductive rights.

*How this study might affect research, practice, or policy:* This study highlights the importance of integrating gender-transformative activities into post-pregnancy contraceptive strategies. It underscores the necessity of understanding and addressing local gender norms and the broader health system context to effectively promote bodily autonomy. The findings suggest that success should not be solely measured by contraceptive uptake but also by how well interventions address gender-related barriers. Future research should focus on developing and validating indicators that evaluate these barriers and promote bodily autonomy, ensuring comprehensive strategies that truly empower women, girls, and gender-diverse individuals with the means, abilities, and assets to make informed choices that resonate with the broader spectrum of their lives.

## INTRODUCTION

Within a framework of sexual and reproductive health (SRH) and rights, the World Health Organization (WHO) aims to fortify the gender-responsive approaches in the escalation and preservation of post-pregnancy contraceptive services that contribute to a fulfilling fertility desire (1). This endeavor stems from recognizing that reproductive and sexual freedom is foundational to gender equality and that “any individual has the necessary agency to determine their reproductive decisions and sexuality, regardless of gendered norms and gender-based discrimination” (2). Such freedom paves the way for women, girls, and those of diverse gender identities to gain comprehensive control over their bodies and life trajectories, irrespective of prevailing restrictive gender perceptions related to sexuality, reproduction, or any gender-related biases. Consequently, the availability of gender-responsive contraception, which goes beyond merely aiming to improve uptake or access to contraceptives but more ambitiously aims to promote personal bodily sovereignty, becomes indispensable in our quest for gender equality.

Accordingly, our study aims consist of evaluating if and how the structure and provision of postpartum and post-abortion contraceptive services empower women, girls, and gender-diverse individuals with the means, abilities, and assets to make informed choices that resonate with the broader spectrum of their lives and challenges harmful gender norms, which may restrict reproductive and life choices. We focus on discerning how such interventions can effectively and intentionally foster bodily autonomy as a primary goal. The evidence generated through this review will advance the knowledge of strategies that successfully address gender-related barriers to scaling up and sustaining postpartum and post-abortion contraception in ways that promote the exercise of bodily autonomy.

## METHODOLOGY

This scoping review follows the Preferred Reporting Items for Systematic Reviews and Meta-Analyses (PRISMA) Statement (3), the PRISMA Extension for Scoping Reviews guidelines (4), and an adaptation of the Arksey and O’Malley framework for conducting scoping reviews (5). The scoping review and its protocol is registered in the OSF repository (6). The guidelines include several stages:

### 1. Identifying the research question

We searched for evidence of addressing gender-related barriers and promoting bodily autonomy in successful strategies to scale up and sustain postpartum and post-abortion contraception. In the absence of other measures in most impact evaluations, we defined success according to the metrics defined for most interventions, that is, increased contraceptive uptake post-pregnancy, therefore, increasing access to sexual and reproductive health services.

### 2. Identifying relevant studies

We searched PubMed, EBSCOhost (Academic Search Complete), and EMBASE online databases. Search terms “in any field” included: ((postpartum contraception) OR (postpartum family planning) OR (post abortion contraception) OR (post abortion family planning) OR (prenatal contraception) OR (prenatal family planning) OR (antenatal contraception) OR (antenatal family planning)) AND ((gender barrier) OR (gender issues) OR (cultural issues)). We also searched the database SciSpace, asking: “What are successful strategies for expanding postpartum or post-abortion contraception?” We included implementation, evaluation, and experimental studies published in any language between 2013 and 2023. We excluded papers that did not meet these criteria. One researcher (AC) conducted all the searches and the following steps in October 2023.

### 3. Study selection

We refined the research strategy in a set of steps. First, we imported all the references to Endnote 20 and deleted duplicates. Second, we imported the remaining references to Covidence and conducted title and abstract screening. We removed publications that were either irrelevant, not found, abstract only, editorials, or letters. Third, we imported the full version of the remaining references plus additional references found through citation search to SciSpace and asked: “Does the paper identify successes to postpartum or post-abortion contraception?” If positive, we refined the search by asking two follow-up questions tested multiple times to capture as many eligible papers as possible: “Were any successes identified?” and “Was the intervention successful?” We excluded the papers that yielded a negative response. We selected the remaining papers for full review and classified them for full review according to the type of studies. We retained implementation, evaluation, and experimental studies and discarded reviews and observational studies. We assessed as eligible the publications that have successfully addressed barriers to the broad adoption and ongoing use of post-pregnancy contraceptive methods, emphasizing the reinforcement of bodily autonomy. One researcher (AC) collected and reviewed the data.

### 4. Charting the data

For each publication, we extracted information on the country where the research was conducted, the population studied, and the methodological design. We charted the data according to four themes:

1. The level of gender responsiveness. To determine the level of each study, we used WHO’s Gender Assessment Tool and Gender Responsive Assessment Scale (7) and additional considerations specific to sexual and reproductive health (2), shown in **Box 1**.
2. Characteristics of gender-responsive strategies that reported success in expanding post-pregnancy contraception, especially those for which impact evaluations have been conducted.
3. Key components of gender-transformative strategies that can be adapted to different contexts and/or scaled up.
4. Factors associated with using digital tools to promote post-pregnancy contraceptive use.

#### Box 1

Levels of gender responsiveness according to the WHO’s Gender Responsive Assessment Scale and gender responsiveness of post-pregnancy contraception services

| Gender Responsive Assessment Scale |  |  |  | Gender responsiveness of post-pregnancy contraception programs |
| --- | --- | --- | --- | --- |
| Level 1 | Gender-unequal | Perpetuates gender inequality by reinforcing unbalanced norms, roles and relations and often leads to one sex enjoying more rights or opportunities than the other. | Hinder the achievement of gender equality and health equity. | Hinder the achievement of sexual and reproductive health and rights. |
| Level 2 | Gender-blind | Ignores gender norms, roles and relations and very often reinforces gender-based discrimination. By ignoring differences in opportunities and resource allocation for women and men, such policies are often assumed to be "fair" as they claim to treat everyone the same. |  |  |
| Level 3 | Gender-sensitive | Indicates gender awareness, although no remedial action is developed. | Represents a starting point for gender programming. | Designed to increase access to adequate post-pregnancy contraception without addressing gender norms and relations. Results are only based on service delivery and specific health outcomes without considering autonomy or informed decision-making. |
| Level 4 | Gender-specific | Considers women's and men's specific needs and intentionally targets and benefits a specific group of women or men to achieve certain policy or program goals or meet certain needs. Such policies often make it easier for women and men to fulfill duties that are ascribed to them based on their gender roles but do not address underlying causes of gender differences. | Represent characteristic s of gender-responsive health policies and programs. | Designed to increase access to appropriate sexual and reproductive health resources through the provision of post-pregnancy contraception. They address gender norms and relations and/or promote informed decision-making. |
| Level 5 | Gender-transformative | Addresses the causes of gender-based health inequities by including ways to transform harmful gender norms, roles, and relations. The objective of such programs is often to promote gender equality and foster progressive changes in power relationships between women and men. |  | Designed to promote empowering reproductive and sexual agency and bodily autonomy through post-pregnancy contraception by transforming gender norms and relations, including those related to the relationship between sexuality and reproduction, addressing gender-based discrimination, and promoting informed decision-making. |

*Source:* Prepared by the authors based on WHO's Gender Responsive Assessment Scale (7) and Coates A, Allotey P. Global health, sexual and reproductive health and rights, and gender: square pegs, round holes. *BMJ Glob Health.* 2023;8(1) (2).

We used the acronyms shown in **Box 2**.

#### Box 2

List of acronyms

|  |  |
| --- | --- |
| ANC | Antenatal care |
| BCS | Balanced Counseling Strategy |
| CHW | Community health worker |
| FIGO | International Federation of Gynecology and Obstetrics |
| FP | Family planning |
| IPP | Immediate postpartum |
| IMCH | Integrated Maternal and Child Health |
| IUD | Intrauterine device |
| L&D | Labor and delivery |
| LAM | Lactation amenorrhea method |
| LARC | Long-acting reversible contraceptive |
| MCH | Maternal and child health |
| MOH | Ministry of Health |
| NGO | Nongovernmental organization |
| PAC | Postabortion care |
| PAFP | Postabortion family planning |
| PNC | Postnatal care |
| PPFP | Postpartum family planning |
| PPIUD | Postpartum intrauterine device |
| RMNCH | Reproductive, maternal, newborn, and child health |
| SRH | Sexual and reproductive health |
| WHO | World Health Organization |

### 5. Patient and Public Involvement

There was no patient or public involvement in this study.

## RESULTS

The PRISMA flow chart (**Figure 1**) shows the identification, screening, and inclusion process of the 30 studies retained. Twenty-seven studies were conducted in a single country and 3 in multiple countries. Combined, the studies were conducted in: Afghanistan (1), Bangladesh (1), Benin (1), Chad (2), China (1), Côte d’Ivoire (1), the Democratic Republic of the Congo (1), Djibouti (1), Ethiopia (3), Ghana (2), Kenya (2), Malawi (1), Mali (1), Nepal (3), Niger (1), Nigeria (2), Pakistan (1), Rwanda (2), Senegal (1), Somalia (1), South Africa (1), Spain (1), Sri Lanka (1), Tanzania (3), Togo (1), and United States (4). All the studies were published in English.

**Figure 1:**
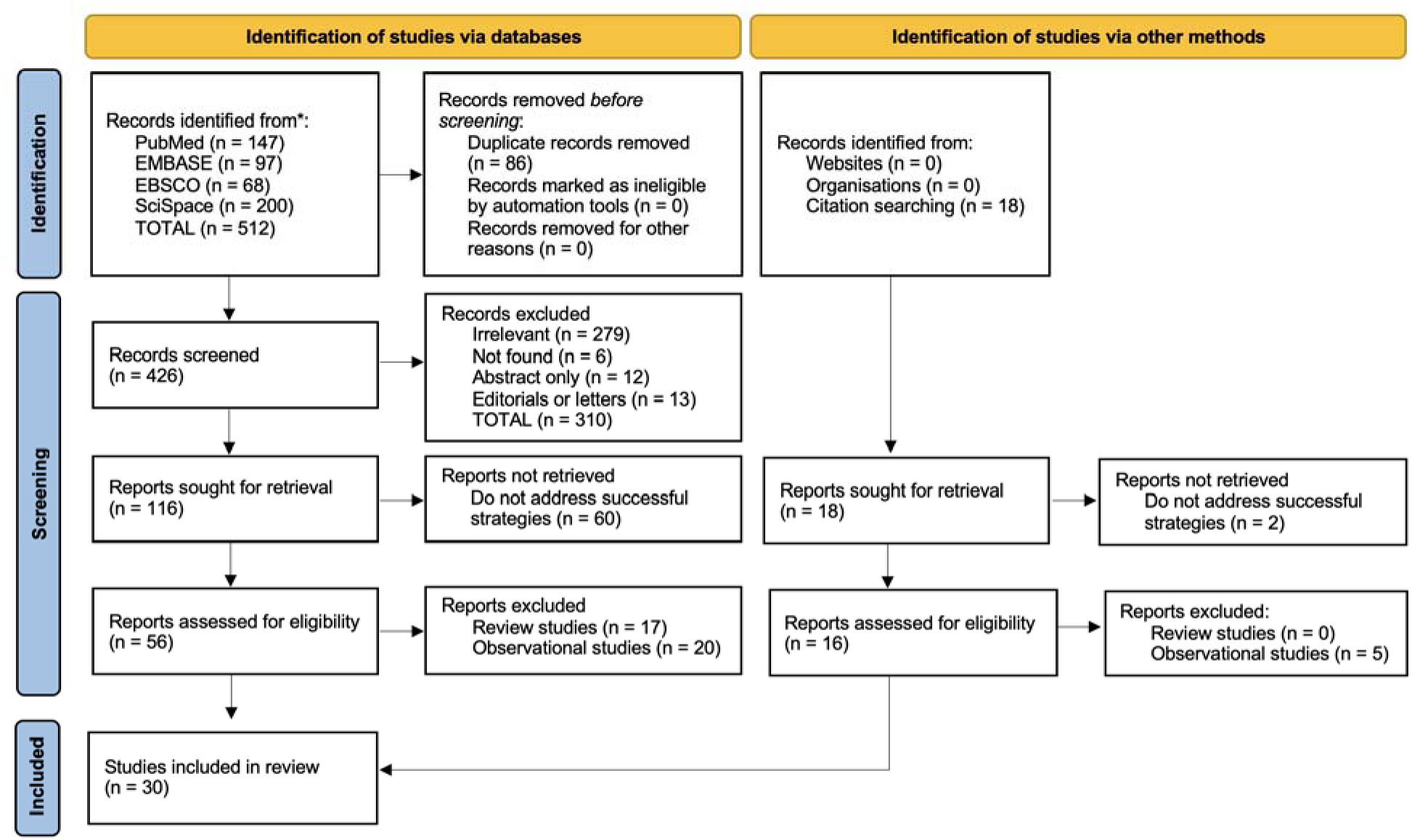
PRISMA flow diagram *Source*: Prepared by the authors following the PRISMA 2020 statement (3).

### 1. Level of gender responsiveness

In

Table 1, we present the 30 studies according to the level of gender responsiveness reported, starting with level 5, in chronological order of publication. Four studies reported gender-transformative strategies (level 5), 24 were gender-specific (level 4), and two were gender-sensitive (level 3). Therefore, none of the strategies hindered the achievement of reproductive health and rights. All the strategies included women and girls, and none were explicitly targeted at gender-diverse people capable of childbearing. Additional information for each study is available in **Annex 1**.

**Table 1:**
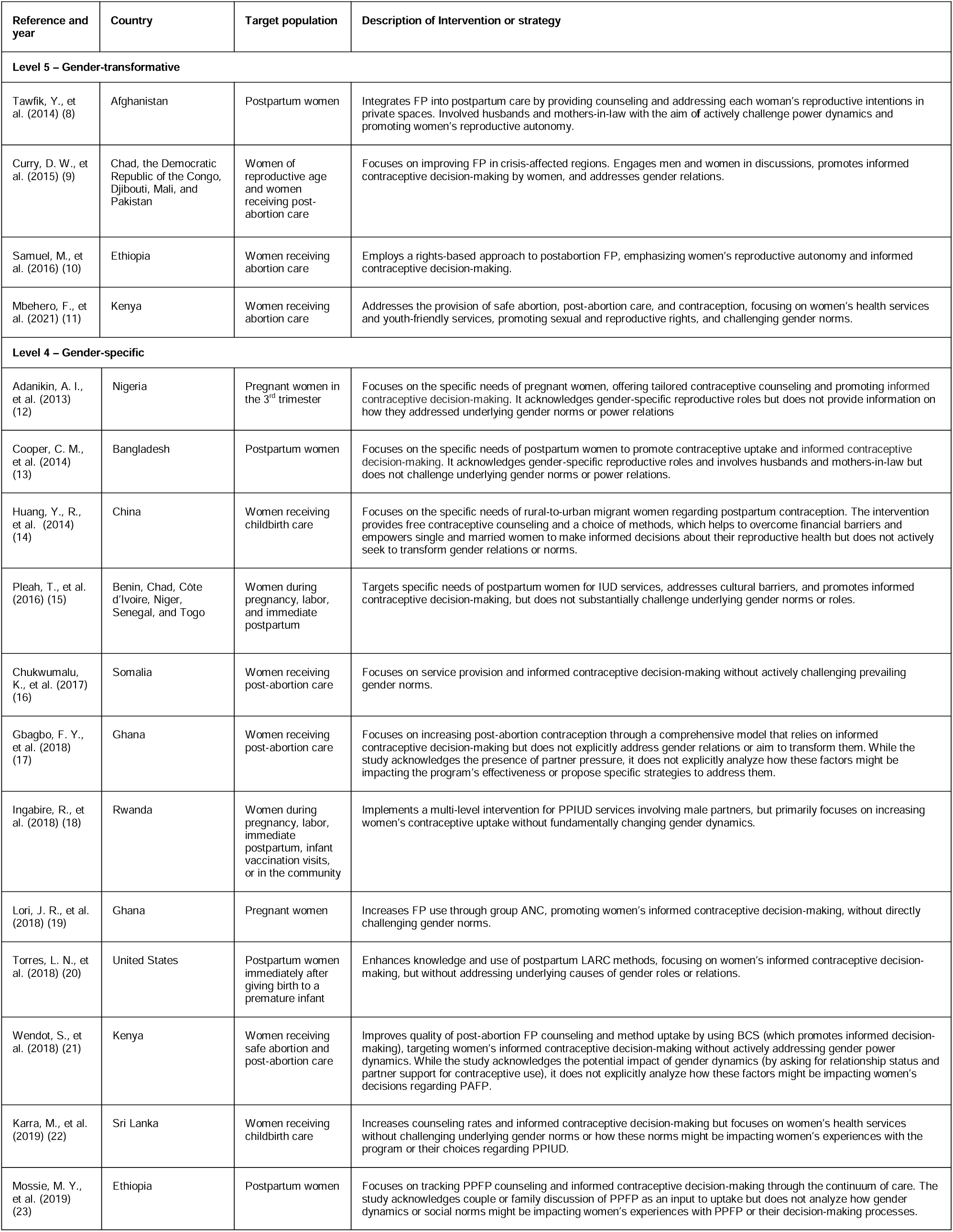
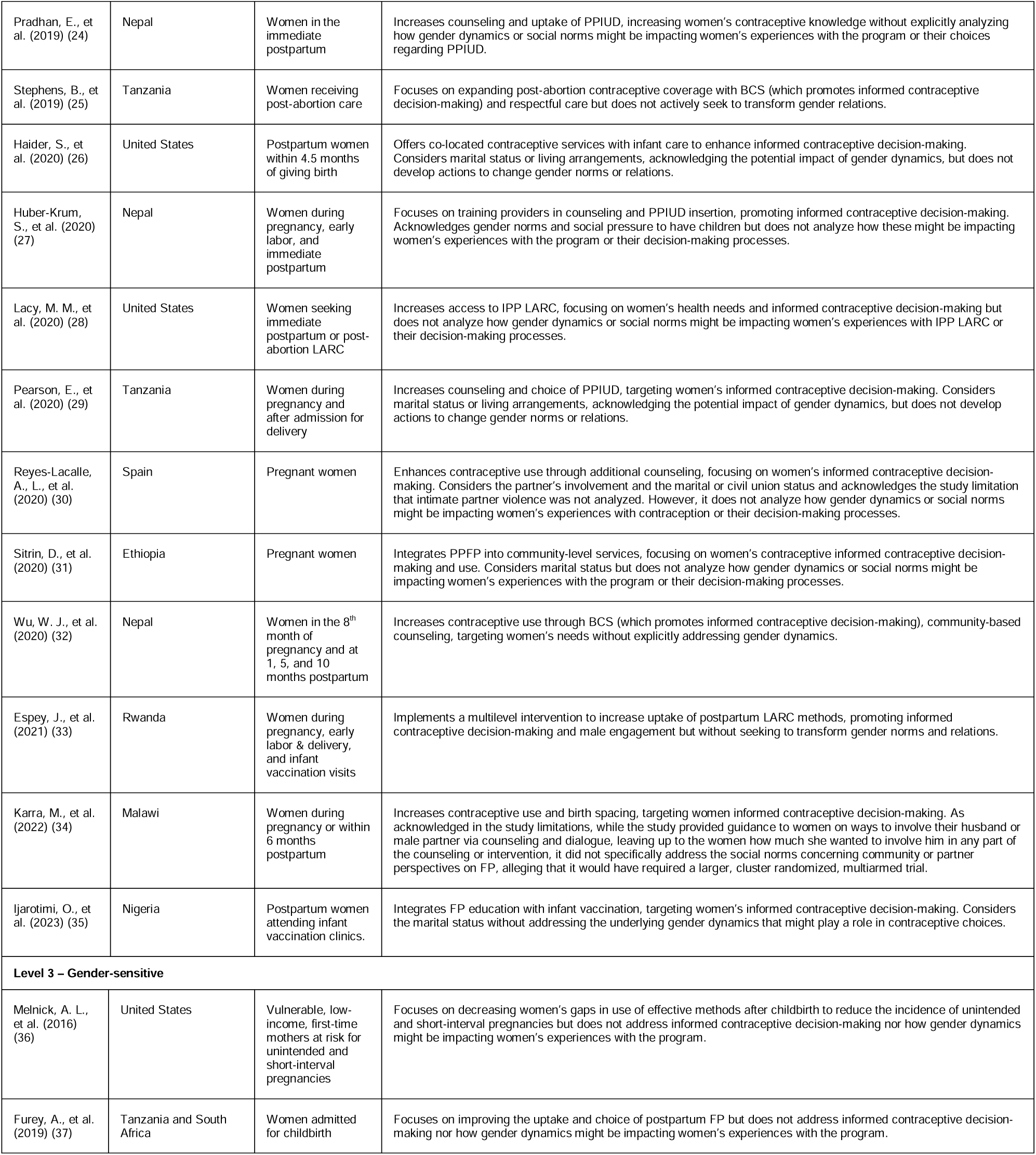
Level of gender-responsiveness of studies that report successful strategies in expanding post-pregnancy contraception.

### 2. Characteristics of gender-responsive strategies that reported success in expanding post-pregnancy contraception

The studies that reported gender-transformative strategies successful in expanding post-pregnancy contraception (8-11) shared the goal of improving the quality of contraception counseling and service provision. These interventions were delivered in contexts of quality improvement strategies that integrated contraception services into the continuum of post-pregnancy care and that offered women continuous access to information. In Afghanistan (8), the intervention involved integrating FP into postpartum care using quality improvement approaches focused on improving the quality of counseling and turning the process user-centered. This included tailoring the counseling process to address each woman’s specific reproductive intentions and contraceptive needs, finding private spaces for PPFP counseling, and involving husbands and mothers-in-law in counseling. Follow-up calls from FP counselors clarified contraceptive methods and underscored essential FP messages. These enhanced approaches successfully integrated FP into postpartum care, resulting in more women receiving counseling on postpartum FP and acquiring their chosen method before leaving the hospital. The strategy addressed gender-based factors by involving husbands and mothers-in-law in FP counseling. Before the strategy, the implementation team had identified that husbands were not allowed in the maternity ward and that some women wanted to consult with their husbands or mothers-in-law before making their decision on contraception use. Creating private spaces for postpartum FP counseling where the husband or the mother-in-law could participate, in person or via mobile phone, was a gender-transformative approach that contributed to meeting women’s needs and a rise in joint decision-making on SRH needs.

In Chad, the Democratic Republic of the Congo, Djibouti, Mali, and Pakistan (9), the choice of contraceptives among women of reproductive age and those receiving post-abortion care varied by country, with LARCs being more prevalent in specific regions and other methods like oral contraceptives and injectables being dominant in others. The study discussed the role of community norms related to gender and fertility in implementing FP services. It emphasized the importance of engaging men and women in discussions about rights, fertility, and contraception at the local level. The strategy also highlighted the role of religious leaders in promoting modern contraception, indicating an understanding of the gender dynamics within religious contexts. Improving FP accessibility and education in crisis-affected regions involved several components: a) facility assessments and stocking: conducting health facility assessments to identify infrastructure gaps, reviewing the presence and condition of basic needs, and reequipping facilities with essential supplies and equipment; b) competency-based training: providing comprehensive training to health care providers, using materials from Jhpiego and the Population Council; this includes both theoretical sessions and practical training; c) supervision and quality improvement: engaging with facility teams and community leaders to ensure a clean, efficient, and respectful environment and regular supervisory visits to assess clinic conditions, stock levels, and data consistency; d) supply chain management: ensuring a steady supply of contraceptive methods and related items by both procuring necessary items and working to strengthen supply chain management in collaboration with government systems; and e) community mobilization: raising awareness at the community level through various means such as radio, participatory theater, and group dialogues, and engaging religious leaders across faiths to raise awareness and ensure women’s rights to access health services. The key to community mobilization’s success was motivating and involving the entire community to recognize the advantages and possibilities in planning and spacing childbirth and facilitating access for women to utilize services.

In Ethiopia (10), the strategy consisted of a package of interventions for women seeking abortion care that promoted a rights-based approach to informed consent and decision-making. The strategy improved the quality of PAFP counseling, increased the contraceptive method mix, emphasized and prioritized respect for women’s reproductive autonomy in the provision of PAFP services, promoted youth-friendly services, ensured providers’ commitment to the sexual and reproductive rights of young women, promoted privacy and confidentiality during service delivery, and strengthened community outreach through health extension workers. These strategies resulted in an increase in postabortion choice of a contraceptive method and a greater proportion choosing the more effective, long-acting methods.

In Kenya (11), health providers were trained on abortion values clarification, attitude transformation, methods of uterine evacuation, management of abortion complications, and post-abortion contraception. Training and mentorship of providers on PAFP and youth-friendly services, along with community engagement and referrals, helped remove common barriers to care: a) the project addressed the issue of providers’ hesitation to counsel and administer LARCs to adolescent or youth users through training and mentoring; b) each facility had separate procedure rooms for conducting medical vacuum aspiration to ensure adequate space and user confidentiality; c) implemented training and mentorship programs for mid-level providers on safe abortion, post-abortion care, and post-abortion contraception, with a focus on youth-friendly services; d) implemented a quality assessment tool based on the WHO’s seven pillars of health systems strengthening and the structures and processes in place to support the domains of quality (safety, timeliness, effectiveness, efficiency, equity, and people-centeredness); e) created health facility registers to track post-abortion contraceptive uptake; f) monitored mentees’ clinical competencies through logbooks, tracking their proficiency in LARC insertion and removal and manual vacuum aspiration; g) monitored the increase in couple counseling for SRH, indicating a rise in joint decision-making on SRH needs; h) community engagement and referral activities to raise awareness and reduce the stigma associated with abortion and contraception; i) incorporation of strong SRH rights advocacy partners into the project to build networks and enhance access to contraception and safe abortion; j) prioritization of youth-led SRH rights advocacy groups in projects to empower young people to make informed decisions about their sexual and reproductive health; k) establishment of quality improvement teams at each facility to assess progress and address gaps, fostering a culture of quality and motivation; l) education of teachers on SRH rights, recognizing the impact of teachers on students’ contraceptive use; and m) implemented strategies to increase male involvement in SRH rights, including holding forums for men and mentoring male champions. The increase in couple counseling for SRH indicated a rising normalization of joint decision-making on SRH needs.

The gender-specific strategies (12-35) focused on informed contraceptive decision-making while recognizing the potential impact of gender dynamics on contraceptive use without actively challenging the underlying gender norms or power relations. The most common gender-specific strategies to expand post-pregnancy contraception included: a) enhancing provider-patient interactions through tailored, structured, or balanced counseling, such as in Nigeria (12), the United States (20, 28), Kenya (21), Tanzania (25); b) increasing the variety of contraceptive choices to better respond to women’s needs, such as in China (14) and Somalia (16); c) integrating contraceptive education with health services like antenatal visits, such as in Ethiopia where they used a modified Integrated Maternal and Child Health to record the woman’s method choice during those visits (23), Nepal (24, 27, 32), Spain (30), Ethiopia (31), Rwanda (33), and Malawi (34); right after admission for delivery services, such as in Tanzania (29); during infant vaccinations, such as in Rwanda (18), Ghana (19), and Nigeria (35); during the first year postpartum, such as in China (14); and other maternal and child health services, such as in the United States (20, 26); d) introducing competency-based training for healthcare providers, such as in Benin, Chad, Côte d’Ivoire, Niger, Senegal, and Togo (15), Somalia (16), Ghana (17); e) employing storytelling with relatable content, such as in Bangladesh (13) and Ghana (19) or showing informational videos in hospital waiting areas, such as in Sri Lanka (22) and Nepal (24). Community engagement, particularly involving religious and community leaders, was critical in addressing cultural and gender norms, as shown in Bangladesh (13), Benin, Chad, Côte d’Ivoire, Niger, Senegal, Togo (15), and Somalia (16).

Finally, the gender-sensitive strategies reported in one study conducted in the United States (36) and one in Tanzania and South Africa (37) were aimed at either increasing the uptake and choice of post-partum contraception through education and training of midwives, nurses, and non-specialist doctors on counseling and provision or by enhancing an existing nurse home visiting program. These studies did not address informed contraceptive decision-making nor how gender dynamics might be impacting women’s experiences with the program.

Among the studies reviewed, we did not find indicators that measure and evaluate gender-related barriers and promote bodily autonomy. **Annex 1** shows a list of indicators based on the strategies that measure and evaluate the rights-based counseling and provision of post-pregnancy contraception.

### 3. Key components of gender-transformative strategies that can be adapted to different contexts or scaled up

Based on the four gender-transformative studies (8-11), **Table 2** shows the multifaceted approach needed to successfully implement gender-transformative interventions in post-pregnancy contraception across different cultural and regional contexts. Details from all the studies are available in **Annex 2**.

**Table 2:** Key components of gender-transformative strategies that can be adapted to different contexts or scaled up.

|  |
| --- |
| **A. Training, Capacity Building, Continuity of Care, and Supply Chain Management** |
| 1. Training healthcare staff in user-centered and rights-based contraception counseling during pregnancy and post-pregnancy to ensure informed decision-making and improve service delivery.<br>2. Training and mentorship programs for healthcare providers on safe abortion, post-abortion care, and contraception, with a focus on youth-friendly services, to sustain quality improvement strategies that support gender-transformative strategies.<br>3. Monitoring and tracking the clinical competencies of providers providing gender-transformative care, e.g. through logbooks.<br>4. Creating private spaces for counseling to ensure privacy and comfort.<br>5. Ensuring follow-up to address each woman's specific reproductive intentions and contraceptive needs, e.g. through follow up calls to answer questions about contraceptive methods.<br>6. Implementing supply chain management approaches that consider local challenges to sustain quality improvement plans that support gender-transformative strategies.<br>7. Monitoring of health systems quality issues, including procurement and supply chains, to ensure women have access to a wide range of contraceptive methods to increase the proportion of post-pregnancy women leaving the facility with their method of choice. |
| **B. Counseling and Empowerment** |
| 1. Tailoring the counseling process to address each woman's specific reproductive intentions and contraceptive needs and prioritize autonomous decision-making.<br>2. Involving key family members, such as partners and mothers-in-law, in counseling sessions to challenge existing power dynamics and promote women's reproductive autonomy and equality in reproductive health decision-making.<br>3. Increasing couple's counseling for SRH to normalize joint decision-making on SRH needs. |
| **C. Community Engagement, Mobilization, and Advocacy** |
| 1. Engaging men and women in local discussions about rights, fertility, and contraception to address and transform cultural and gender-based norms and relations.<br>2. Community mobilization, engagement, and referral activities through radio, participatory theater, and group dialogues, involving religious leaders across faiths and local leaders to discuss the role of community norms related to gender and fertility and raise awareness and support women's rights to access health services.<br>3. Increasing male involvement in discussions about sexual and reproductive health and rights, fertility, and contraception, including holding forums for men and mentoring male champions, to address and transform cultural and gender-based norms and relations at the local level.<br>4. Prioritization of youth-led SRH rights advocacy groups in projects to address and transform cultural and gender-based norms and relations.<br>5. Incorporation of strong SRH rights advocacy partners in the interventions to build networks and enhance access to contraception and safe abortion.<br>6. Engaging in dialogue with local community groups, especially women's organizations to better understand local gender norms and women's reproductive needs. |

*Note:* Prepared by the authors based on gender-transformative studies (8-11).

### 4. Factors associated with using digital tools to promote post-pregnancy contraceptive use

Of the 30 studies included in the review, ten referred to the use of digital tools to promote post-pregnancy contraceptive use (15, 18, 23, 25-30, 32). Two studies conducted in Nepal and one in Spain reported that digital technology in healthcare improved service integration and facilitated the dissemination of contraceptive information (27, 30, 32). In Tanzania, mobile applications and digital counseling tools were instrumental in training healthcare workers, giving them access to the latest information and techniques to meet users’ needs effectively (25). Digital data collection and management systems enhanced the continuity and follow-up of women users of contraception. For example, in Benin, Chad, Côte d’Ivoire, Niger, Senegal, and Togo, digital tools helped identify PPIUD complications (15). In Ethiopia, digital health records helped improve service integration and continuity of care, track PPFP, and facilitate decision-making and timely adoption of contraceptives (23). In Spain, women received reminders to consult contraception information during pregnancy (30). In Nepal, community health workers used mobile applications to individualize the counseling based on women’s responses to the balanced counseling strategy questions and to follow up at subsequent visits on initiation, barriers to access, and continuation (32). Finally, tablets and software for data input were used to collect data in Rwanda (18), Tanzania (29), the United States (26, 28), Spain (30), and Nepal (32).

## DISCUSSION

We reviewed 30 studies that met all the inclusion criteria. All but two studies emphasized the need to provide a supportive environment that respects and enhances women’s bodily autonomy. The gender-transformative strategies reported in four of the studies shared a commitment to empowering women with the autonomy to make informed decisions about post-pregnancy contraception. They achieved this through quality improvement strategies that seamlessly integrated contraception services into the continuum of postpartum and post-abortion care. Providers delivered personalized counseling that respected each woman’s reproductive goals and ensured privacy during these discussions. The initiatives included follow-up support to offer women continuous access to information and reinforce contraceptive practices. These efforts promoted a rights-based approach, prioritizing informed consent and defending women’s reproductive rights. Women often chose long-acting reversible contraceptives, reflecting a preference for more reliable and enduring contraception methods when they received comprehensive information. These programs acknowledged and integrated the cultural and family context of women’s health decisions by involving husbands and mothers-in-law, not as vehicles to increase post-pregnancy contraception but to challenge existing power dynamics and promote women’s reproductive autonomy and equality in reproductive health decision-making. The implementation of these strategies, carefully adapted to each context, underscores the effectiveness of a comprehensive, multi-layered approach in transforming health interventions related to gender dynamics. By tackling both service delivery and broader social elements like gender norms and community standards, the findings from the gender-transformative strategies illustrate the potential for significant advancements in expanding post-pregnancy contraception.

The gender-specific studies focused on informed contraceptive decision-making. The studies demonstrated that multiple counseling sessions during antenatal care, tailored counseling during the third trimester, the free provision of contraception integrated into childbirth and abortion care, a range of options in contraceptive methods, and follow-up visits were crucial in ensuring the informed contraceptive decision-making process. Training healthcare providers in insertion techniques and providing informed consent and rights-based counseling was pivotal in increasing the informed uptake of post-pregnancy LARC methods and, specifically, PPIUD. Community engagement strategies that included the participation of religious and community leaders were instrumental in facilitating women’s access to health facilities for post-abortion care services. The strategies also acknowledged the need for comprehensive monitoring and evaluation frameworks for data-driven programmatic decisions, engagement of key stakeholders from the initial stages, and the importance of youth-friendly services.

While they recognized the potential impact of gender dynamics on contraceptive use and some interventions involved key family members in the counseling process, some strategies did not actively challenge the underlying gender norms or power relations.

Although several scoping and systematic reviews have addressed strategies that increase post-pregnancy uptake of contraception around the world (38-46), this is the first scoping review of interventions that promote bodily autonomy when addressing gender-related barriers to scaling up and sustaining post-pregnancy contraception.

The review has allowed us to identify key elements of gender-transformative post-pregnancy contraceptive strategies, with some important components that can be adapted to diverse contexts.

Personal counseling emerged as a common thread across many interventions within a framework of a user-centered approach. Similarly, community dialogue and involvement of advocacy organizations, especially women’s organizations, are crucial to understanding local gender norms and women’s reproductive needs. Both are fundamental to delivering the individualized care required to promote bodily autonomy. However, for bodily autonomy to be meaningfully realized, other elements are also vital. With some notable exceptions, few strategies noted the wider health system context, such as procurement and supply chains to enable providers to meet women’s contraceptive needs, institutionalized quality assurance mechanisms stressing gender transformative approaches, and ensuring an enabling health policy and legal environment.

Understanding cultural and familial contexts is another key element. The precise intent of strategies tailored to context is significant to ensure that they do not inadvertently reinforce unequal power relations, rather than advancing women’s bodily autonomy. It is, therefore, especially significant that although the review allowed us to generate a list of results indicators to measure and evaluate the rights-based counseling and provision of post-pregnancy contraception, we did not find indicators that measure and evaluate gender-related barriers and promote bodily autonomy. Whilst success is still only defined according to contraceptive access and uptake, it cannot be explicitly guaranteed that any intervention will be delivered in such a manner that it will meaningfully promote bodily autonomy,

We recommend that future research urgently focus on developing and validating indicators that specifically address gender-related barriers and promote bodily autonomy so that contraceptive uptake is not mistakenly understood as a sole proxy for bodily autonomy, but rather as a dual goal. There is a need for more comprehensive strategies that integrate these indicators—and thus approach and goals—into practice, ensuring that post-pregnancy contraception programs empower women and girls, with the tools, knowledge, and resources to make informed decisions that align with their life goals.

## Data Availability

All data produced in the present work are contained in the manuscript. Additional data will be available when the manuscript is published.

